# Understanding the Biering-Sørensen test: contributors to extensor endurance in young adults with and without low back pain

**DOI:** 10.1101/2023.01.11.23284452

**Authors:** Jonathan Shaw, Jesse V. Jacobs, Linda R. Van Dillen, George J. Beneck, Jo Armour Smith

**Affiliations:** Chapman University; Rehabilitation and Movement Science, University of Vermont; Program in Physical Therapy, Orthopaedic Surgery, Washington University School of Medicine; Department of Physical Therapy, California State University, Long Beach; Department of Physical Therapy, Chapman University

**Keywords:** Biering-Sørensen test, low back pain, endurance, paraspinal

## Abstract

Impaired paraspinal muscle endurance may contribute to persistent low back pain (LBP) and is frequently assessed using a single repetition of the Biering-Sørensen test. This study investigated how Sørensen test duration, muscle activation, and muscle fatigability are affected by multiple repetitions of the test, and determined predictors of Sørensen test duration in young, active adults with and without a history of LBP. Sixty-four individuals with and without persistent LBP performed 3 repetitions of the Sørensen test. Amplitude of activation and median frequency slope (fatigability) were calculated for the lumbar and thoracic paraspinals and the hamstrings. Duration of the test was significantly less for the 2^nd^ and 3^rd^ repetitions in individuals with LBP. In individuals without LBP, fatigability of the lumbar paraspinals was the best predictor of test duration. In individuals with LBP, Sørensen test duration was predicted by fatigability of the hamstrings and amplitude of activation of the thoracic and lumbar paraspinals. Our findings demonstrate that it is necessary to amplify the difficulty of the Sørensen test to elucidate impairments in young, active adults with LBP. Training programs aiming to improve lumbar paraspinal performance in individuals with LBP should monitor performance of other synergist muscles during endurance exercise.

## Introduction

Most individuals with persistent low back pain (LBP) have their first episode of pain as an adolescent or young adult.^1^ As episodes of LBP become increasingly disabling with time it is important to establish potential contributors to LBP early in the lifespan.^2^ Impaired paraspinal muscle endurance may contribute to persistent back pain via altered motor control and maladaptive spinal loading.^3–10^

The Biering-Sørensen test is commonly used to assess endurance of the paraspinal musculature using two metrics: test duration (the time that an individual maintains the Sørensen test position), and rate of muscle fatigue (the rate at which the median frequency of the muscle electromyography power spectrum declines over time).^11,12^ Increasing amplitude of muscle activation during the Sørensen test is also an indication of fatigue.^13^ To date, results from research using the Sørensen test to investigate the relationship between endurance and current or future LBP have been mixed.^3,12,14–19^ The conflicting findings across studies may be due the multiple person-specific biopsychosocial characteristics that influence Sørensen test duration and rate of fatigue.^19^ One-such characteristic is level of physical activity. Typical physical activity may influence both duration of the test and muscle fatigability due to deconditioning or muscle disuse atrophy.^16,19^ Psychological factors that may influence test duration, but not EMG measures of fatigability, include generalized fear of movement (fear avoidance^20^), pain catastrophizing,^19^ anxiety,^21^ motivation and fear during the test.^16,22^

The Sørensen test is commonly used as test of the lumbar paraspinal muscles, but the thoracic paraspinals and the hip extensors also contribute to maintenance of the test position.^23,24^ It is not clear how thoracic paraspinal or hip extensor activation or fatigability influence the Sørensen test, in individuals with and without LBP.^5,18,23^ Typically, only a single repetition of the Sørensen test is used. However, in young, active adults, a single repetition may be insufficiently fatiguing to identify muscle performance impairments related to LBP.^5^

The purpose of this study was to determine how Sørensen test duration, muscle activation and fatigability of the lumbar and thoracic paraspinals and hip extensors are affected by multiple repetitions of the Sørensen test in young, active adults with and without a history of LBP. The second purpose of the study was to determine factors that predict the duration of the Sørensen test after multiple fatiguing test repetitions have been performed in young adults with and without a history of LBP.

## Materials and Methods

### Participants

Young adults between the ages of 18 and 35 years participated. Individuals in the LBP group had a history of functionally limiting low back pain of at least one year’s duration. They were in symptom remission at the time of the data collection. The group without LBP included individuals with no significant history of LBP. Exclusion criteria for both groups included inflammatory or neuropathic disease, spinal stenosis, radiculopathy, spondylolisthesis, or scoliosis. Participants gave written consent before participating. Chapman University’s Institutional Review Board approved the study.

Participants completed the Physical Activity Scale (PAS). This validated measure quantifies physical activity over a typical 24-hour period, in metabolic equivalents (METS).^25,26^ Participants with LBP also identified the duration and frequency of their symptoms and completed measures of average pain intensity during symptomatic episodes (visual analogue scale, 0 – 10), the Fear Avoidance Beliefs Questionnaire physical activity subscale (FABQ-P^27^) and the modified Oswestry Disability Index (ODI^28^).

### Instrumentation

Participants were instrumented with surface electromyography electrodes (interelectrode distance 20mm, Myotronics Inc, WA, USA). Following standard skin-preparation procedures,^29^ electrodes were placed bilaterally on the lumbar erector spinae/multifidus (LES, 2cm lateral to the L4/L5 interspace, thoracic erector spinae (longissimus thoracis pars thoracis, TES, lateral to the spinous process of T10 on the muscle belly), and hamstrings (biceps femoris, HS, midway between the ischial tuberosity and the lateral tibial epicondyle),^29–31^ and attached to wireless sensors digitally sampling at 1500Hz (Noraxon DTS sensors, Noraxon Inc, Scottsdale, USA).

### Fatiguing exercise (Biering-Sørensen Test)

Participants performed three Sørensen tests. Between each test, participants performed repeated sub-maximal movements involving either raising a leg in supine, raising an arm in standing, or walking for approximately ten minutes. These tasks have been described elsewhere, and the order in which they were completed was randomized.^32–34^ Because these tasks required different postural orientations and did not involve the low back as the primary mover, we defined these times of activity between Sørensen tests as active-rest periods.

For the Sørensen tests, participants lay on a Roman chair with their lower limbs stabilized and their head, arms, and trunk unsupported (Figure 1). Participants were instructed to maintain a horizontal body position for as long as possible. The correct test position was monitored visually by observing the change in position of a plumb bob hung from the participants’ necks.^16^ Participants were given standardized verbal encouragement. The end of the test occurred when the participant could no longer maintain the horizontal test position or when the participant felt unable to continue the test. After each repetition of the Sørensen test, any back pain/discomfort in the low back area that persisted during the ten-minute active-rest period was quantified using a visual analogue scale.

**Figure 1.**
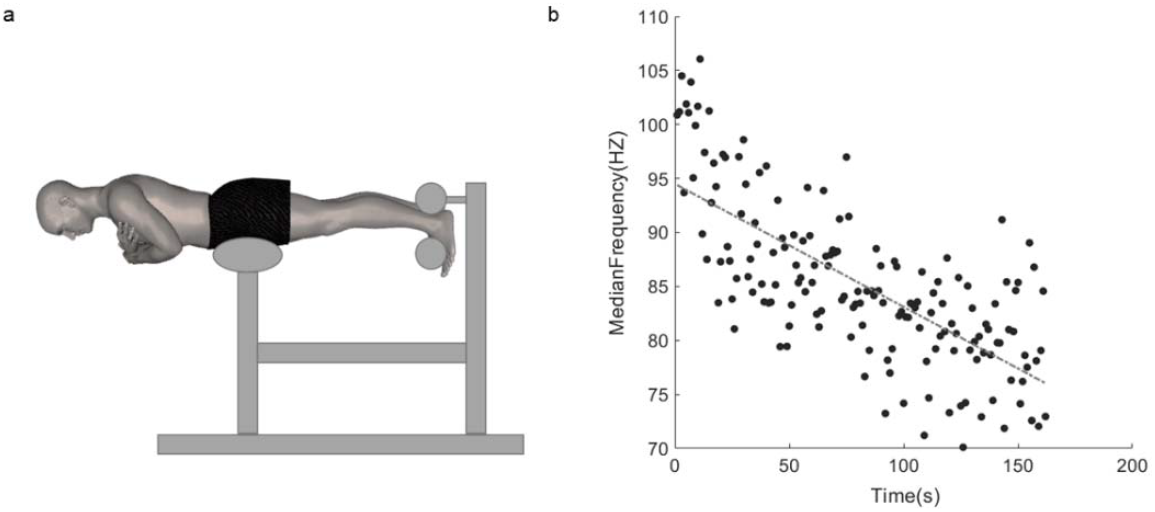
a) Testing position for the Biering-Sørensen test. b) Representative data from the lumbar paraspinal muscle from a single participant showing the decline in the median frequency of the electromyography signal over the duration of a test. Fatigability is quantified as the slope of the decline in median frequency, normalized to the initial median frequency.

### EMG Analysis

EMG data were band-passed filtered (30-450 Hz) and then notch-filtered to remove environmental noise (MATLAB® version R2018B, The MathWorks, Natick, MA). The median frequency of the power spectrum, in Hz, was calculated for every second of the entire Sørensen test using a Fast Fourier transform. The slope of the median frequency values over time was calculated using least squares linear regression.^35,11,16,36^ Median frequency slopes were normalized to the initial median frequency to account for individual differences in subcutaneous tissue depth.^37^ Root mean square amplitude of activation was calculated for every second of the each Sørensen test and then averaged for each repetition. Average amplitude during all three repetitions was normalized to the root mean square average of the first ten seconds of the first Sørensen test.

### Statistical Analysis

Demographics and the Sorensen test characteristics were compared between groups with independent t-tests or chi^2^ tests. Normalized median frequency slope and amplitude were analyzed with separate mixed-model ANOVA tests, testing for main effects of repetition and group and repetition*group interactions. Post hoc comparisons were made with Bonferroni correction (SPSS^®^ Statistics Version 26, IBM^®^, Armonk, USA).

Predictors of test performance were examined using the duration of the final, third Sørensen test. We hypothesized that participants would be most fatigued during the third repetition, and that therefore this repetition would be most sensitive to highlighting differences between groups. Bivariate relationships between the duration of the third Sørensen test repetition, (Duration 3) and potential explanatory variables were tested with Pearson correlation coefficients for each group separately. Variables that were correlated with Duration 3 (p < 0.05) were then entered into separate ordinary least squares regression models using a backward stepwise approach (α enter p = 0.05 and α exit p = 0.10). Models were checked for influential outliers using Cook’s Distance.

## Results

Participant demographics and Sørensen test characteristics are shown in Table 1. There was no difference between the groups in age, distribution of sex, or total and vigorous physical activity. Individuals with LBP had significantly higher BMI (p = 0.004). To control for the effect of this group difference, BMI was mean-centered and entered as a covariate in the ANOVA models.

**Table 1.** Group demographics and Sørensen Test duration (means and standard deviations)

|  | **With LBP**<br>**(n = 41)** | **Without LBP**<br>**(n = 23)** |
| --- | --- | --- |
| Age (years) | 21.9 (3.1) | 23.5 (3.6) |
| Sex |  |  |
| Male | 16 | 9 |
| Female | 24 | 14 |
| Not specified | 1 |  |
| BMI (kg/m<sup>2</sup>) | **24.3 (4.5)** | **21.8 (2.2)** |
| PAS (METs) | 46.9 (11.4) | 45.1 (12.8) |
| PAS-VIG (METs) | 10.1 (13.4) | 7.8 (8.0) |
| Duration 1 (s) | 99.2 (37.6) | 112.5 (44.8) |
| Duration 2 (s) | **89.3 (29.0)** | **109.8 (37.8)** |
| Duration 3 (s) | **80.2 (25.0)** | **101.2 (36.8)** |
| Post-test pain 1 (VAS) | **0.6 (0.7)** | **0.2 (0.4)** |
| Post-test pain 2 (VAS) | **0.7 (0.7)** | **0.3 (0.6)** |
| Post-test pain 3 (VAS) | 0.8 (0.8) | 0.4 (0.8) |
| Typical pain | 4.8 (2.2) | n/a |
| FABQ-P | 9.2 (4.8) | n/a |
| ODI | 16.0 (13.3) | n/a |

BMI – Body Mass Index. PAS – Physical Activity Scale. PAS-VIG – Physical Activity Scale, vigorous sub-scale, measured in metabolic equivalents (METs). VAS – Visual Analogue Scale for Pain, 0 – 10. FABQ-P – Fear Avoidance Beliefs Questionnaire, physical sub-scale. ODI – Modified Oswestry Disability Index. Typical pain is the average pain that participants reported they experienced during symptomatic episodes. Characteristics with significant differences between groups are shown in bold.

All experimental data from one participant without LBP were excluded because the participant reported that the duration of their Sørensen test was limited by anterior thigh discomfort rather than paraspinal or hip extensor fatigue. Two participants with LBP did not complete the third repetition; one because of anxiety about exacerbating their symptoms, and one because of time constraints. All other participants with LBP completed the full testing protocol without difficulty and reported that Sørensen test duration was limited by fatigue rather than reproduction of their symptoms. EMG sensor failure prevented collection of the third repetition from one participant. Visual inspection of EMG signals by the first and senior authors revealed that two or three individuals had EMG signal with periods of signal drop-out for each muscle due to loss of sensor connectivity and these signals were excluded after a consensus decision was reached.

### Group comparisons

Normalized median frequency slopes and normalized amplitude for the right and left sides of all three muscles were significantly correlated (p < 0.05 for all muscles and all repetitions). Therefore, the average of the slope and the average of the amplitude from both sides was calculated for each repetition and these average variables were used in all further analyses (thoracic erector spinae TES1, TES2, TES3; lumbar erector spinae LES1, LES2, LES3; hamstrings HS1, HS2, HS3).

For TES, the normalized median frequency slope declined significantly across the test repetitions (main effect of repetition F = 4.932, p = 0.009), indicating an increasing rate of fatigue. The rate of fatigue was significantly greater for repetitions two and three than repetition one (Bonferroni adjusted p < 0.05 for all comparisons, Figure 2a). For LES median frequency there was a repetition*group interaction (interaction F = 3.341, p = 0.049). For the control group the rate of fatigue increased at each repetition (p < 0.05 for all comparisons, Figure 2b). For the LBP group, the rate of LES fatigue was not significantly different between repetitions 1 and 2 but was significantly greater for repetition 3 (p < 0.05). For the HS, rate of fatigue was significantly greater in the second and third repetitions compared with the first (F = 12.343, p < 0.001; p = 0.024 and p < 0.001 respectively, Figure 2c).

**Figure 2.**
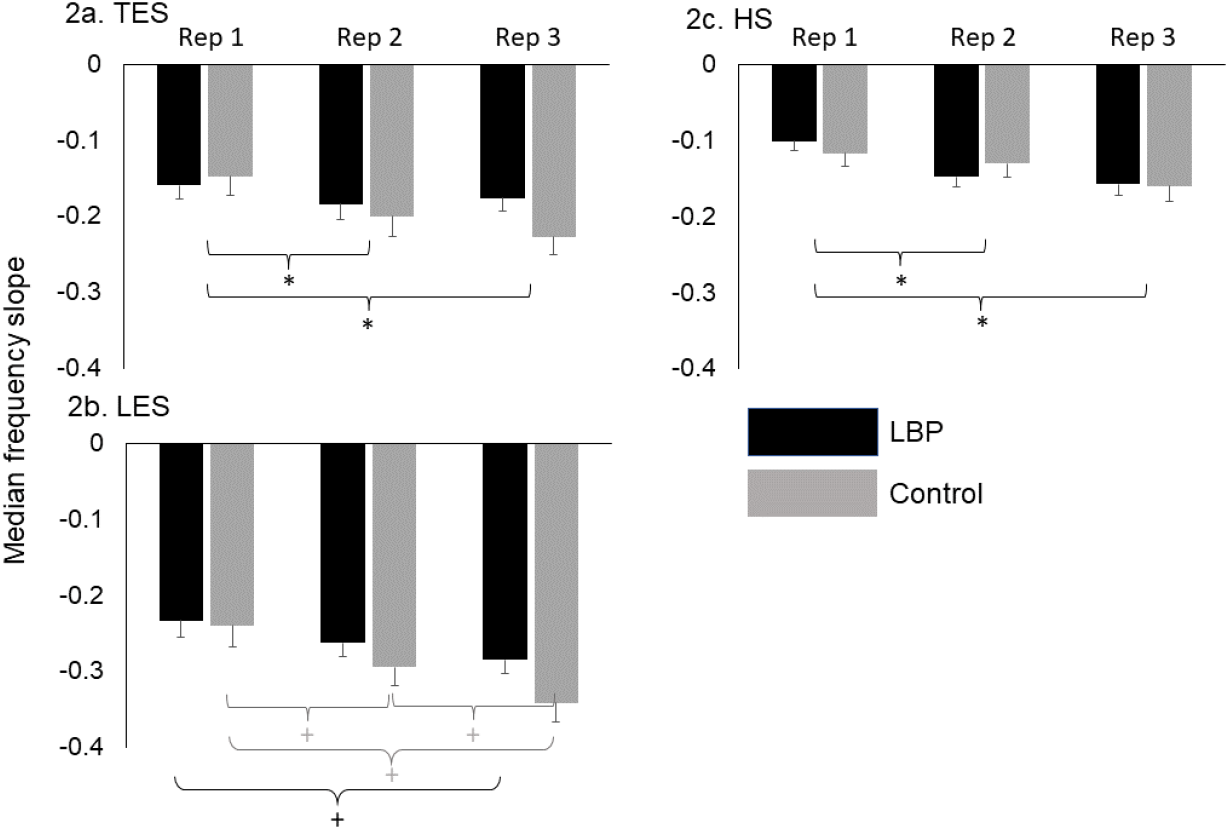
Normalized median frequency slope for a) Thoracic Erector Spinae (TES), b) Lumbar Erector Spinae (LES), c) Hamstring (HS), with significant post-hoc comparisons for the main effect of repetition (Rep 1, 2, 3) indicated by *, and significant within-group post-hoc comparisons indicated by +. N = 37 in the group with LBP and 21 in the group without LBP. Values are marginal means after covarying for BMI and error bars are standard errors.

Normalized amplitude of TES increased significantly across repetitions (F = 59.632, p < 0.001). TES amplitude increased at each repetition (p < 0.001 for all comparisons, Figure 3a). Similarly, amplitude of LES activation increased across repetitions (F = 57.382, p < 0.001, all post-hoc comparisons p < 0.001) but did not differ by group. Amplitude of HS activation also increased across repetitions (F = 26.789, p < 0.001) but there was no effect of group or repetition*group interaction. Normalized amplitude of HS activation increased at each repetition (p < 0.02 for all comparisons).

**Figure 3.**
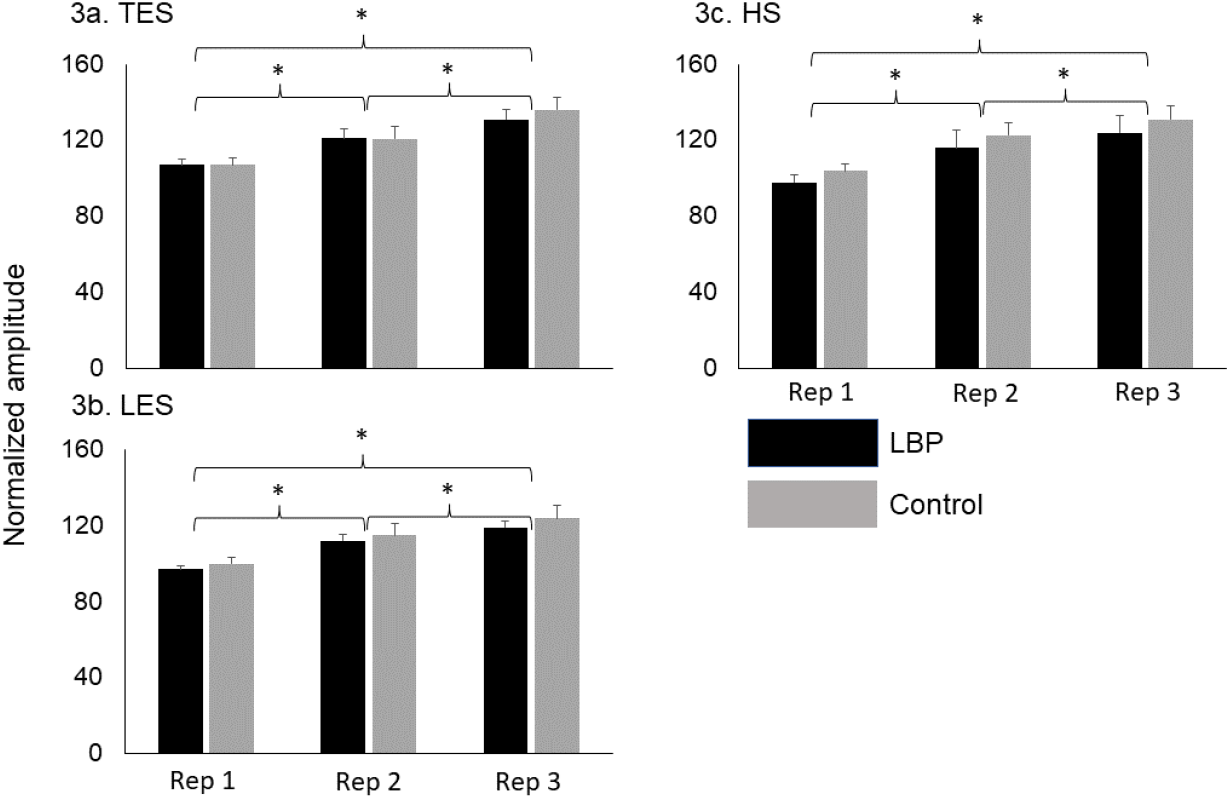
Normalized amplitude for a) Thoracic Erector Spinae (TES), b) Lumbar Erector Spinae (LES), c) Hamstring (HS), with significant post-hoc comparisons for the main effect of repetition (Rep 1, 2, 3) indicated by *. N = 37 in the group with LBP and 21 in the group without LBP. Values are marginal means after covarying for BMI and error bars are standard errors. Data were normalized to average amplitude of activation during the first ten seconds of the first repetition of the Sørensen test.

### Predictors of third test duration

Bivariate relationships between candidate explanatory variables and Duration 3 are shown in Table 2 and Figure 4. In the group with LBP, greater age was associated with shorter Sørensen test duration. The normalized slopes of TES1, LES1, HS1 and HS2 and the normalized amplitude of TES2 and LES3 were also significantly associated with duration. Individuals in the LBP group with faster rate of fatigue (steeper slope) and lower amplitude of activation had shorter Duration 3. In the group without LBP, greater age, and greater rate of fatigue in LES1, LES2, and LES3 were significantly correlated with shorter duration of the third Sørensen repetition. There was no significant correlation between amplitude of activation and duration of the Sørensen test in the group without LBP.

**Table 2.** Linear relationship between duration of the third Sørensen test, in seconds, and potential predictor variables. Note that, for median-frequency slopes, positive correlation indicates that a more negative slope (faster rate of fatigue) was associated with shorter duration of the third Sørensen repetition. For amplitude, positive correlation indicates that greater amplitude of activation was associated with longer duration of the third Sørensen repetition.

|  | LBP | Control |
| --- | --- | --- |
| Age (years) | **-0.318** | **-0.541** |
| BMI ((kg/m<sup>2</sup>)) | -0.125 | -0.269 |
| PAS (METS) | 0.154 | -0.160 |
| PAS-VIG (METS) | 0.015 | -0.260 |
| TES1 (normalized slope) | **0.453** | -0.117 |
| LES1(normalized slope) | **0.412** | **0.471** |
| HS1 (normalized slope) | **0.422** | 0.306 |
| TES2 (normalized slope) | 0.263 | 0.134 |
| LES2 (normalized slope) | 0.197 | **0.512** |
| HS2 (normalized slope) | **0.327** | 0.319 |
| TES3 (normalized slope) | 0.104 | 0.436 |
| LES3 (normalized slope) | 0.246 | **0.649** |
| HS3 (normalized slope) | 0.207 | 0.413 |
| TES1 (normalized amplitude) | 0.149 | -0.166 |
| LES1(normalized amplitude) | -0.206 | -0.176 |
| HS1 (normalized amplitude) | 0.064 | -0.052 |
| TES2 (normalized amplitude) | **0.364** | 0.317 |
| LES2 (normalized amplitude) | 0.259 | -0.255 |
| HS2 (normalized amplitude) | 0.163 | -0.209 |
| TES3 (normalized amplitude) | 0.291 | -0.096 |
| LES3 (normalized amplitude) | **0.370** | -0.239 |
| HS3 (normalized amplitude) | 0.223 | -0.055 |
| Post-test pain 1 (VAS) | 0.177 | 0.086 |
| Post-test pain 2 (VAS) | -0.052 | 0.143 |
| Post-test pain 3 (VAS) | 0.065 | 0.154 |
| Typical pain (VAS) | 0.126 | n/a |
| FABQ-P | 0.127 | n/a |
| ODI | -0.095 | n/a |

BMI – Body Mass Index. PAS – Physical Activity Scale. PAS-VIG – Physical Activity Scale, vigorous sub-scale. VAS – visual analogue scale for pain. FABQ-P – Fear Avoidance Beliefs Questionnaire, physical sub-scale. ODI – Modified Oswestry Disability Index. TES – Thoracic Erector Spinae. LES – Lumbar Erector Spinae. HS – Hamstring. Significant correlations ( $p < 0.05$ ) are shown in bold.

**Figure 4.**
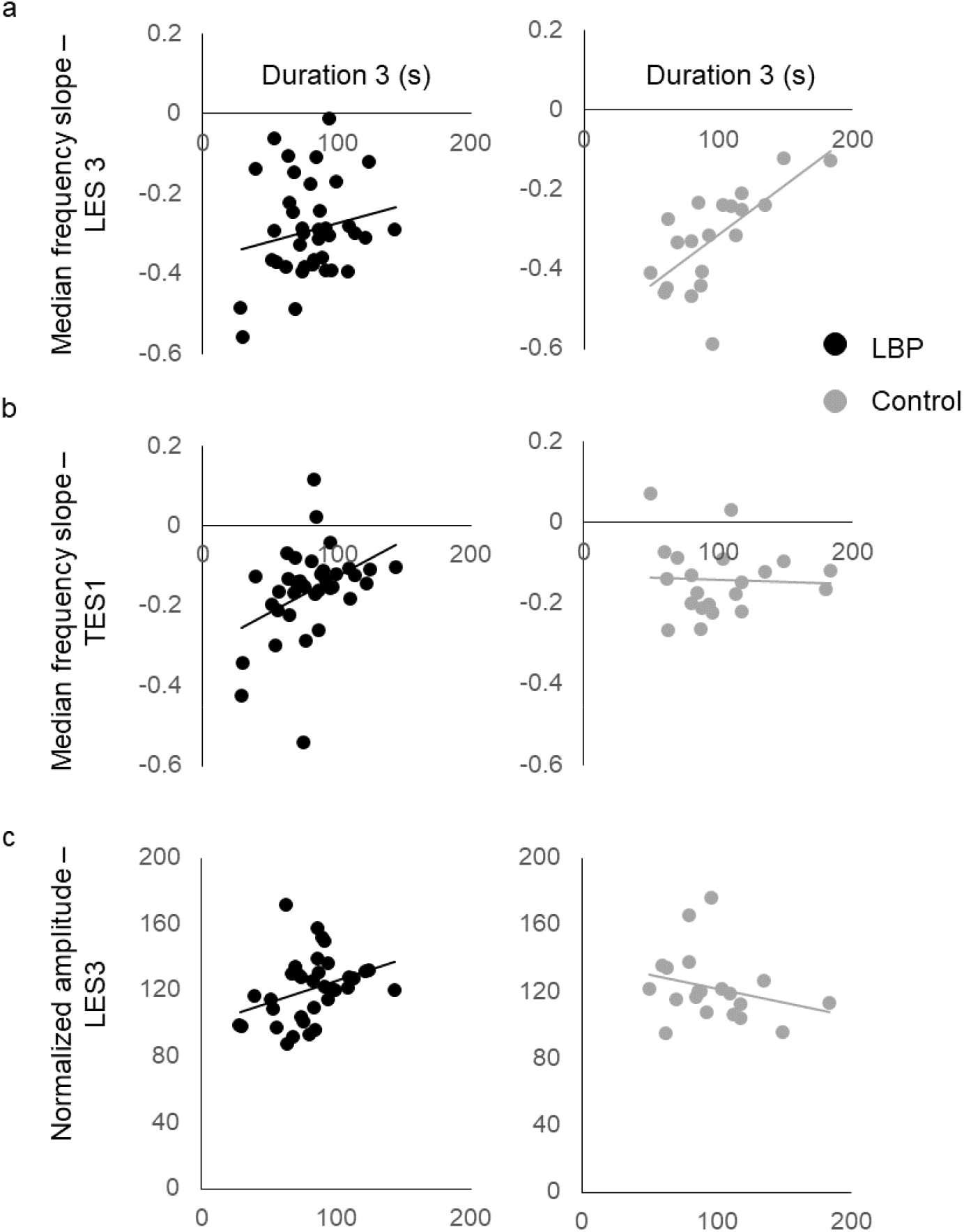
Scatterplots showing relationship between duration of the third Sørensen test in seconds and a) normalized median frequency slope of the lumbar paraspinals during the third test (LES3); b) normalized median frequency slope of the thoracic paraspinals during the first test (TES1); c) normalized amplitude of the lumbar paraspinals during the third test.

The explanatory variables with significant bivariate relationships were entered into the multivariate linear regression models. For the group with LBP, duration of the third Sørensen repetition was predicted in the final model by a combination of HS1 slope, TES2 amplitude and LES3 amplitude (R^2^ = 0.424, F = 7.114, p = 0.001, β HS1 slope 0.407, β TES2 amplitude 0.309, β LES3 amplitude 0.312). For the group without LBP, Duration 3 was predicted solely by the slope of LES3 (R^2^ = 0.652, F = 13.317, p = 0.002, β LES3 = 0.652).

## Discussion

In active young adults with a history of LBP, more than one repetition of the Sørensen test is needed to identify impairment in paraspinal/hip extensor endurance. Despite being minimally disabled by their symptoms, young adults with LBP had significantly poorer test endurance for the second and third test repetitions than individuals with no history of LBP. Physical activity and other person-specific characteristics did not significantly influence the duration of the third repetition of the Sørensen test. In young adults without LBP, the fatigability of the lumbar paraspinals was the most important influence on the duration of the third Sørensen test. However, in individuals with a history of LBP, the duration of the final test was dependent upon the fatigability of the hamstrings, and the extent to which participants activated the thoracic and lumbar paraspinals.

We did not observe reduced test duration in the group with LBP for the first repetition of the Sørensen test. This suggests that the first repetition of the test did not fatigue individuals sufficiently to demonstrate group differences. During any fatiguing exercise test, muscle performance is influenced by multiple mechanisms. These include peripheral muscle structure and function, neural activation strategy, and sense of effort.^38^ During the Sørensen test, the submaximal nature of the test may result in the participant’s perception of fatigue (sense of effort) being the limiting factor rather than decreased ability to generate force.^5,38^ As a result, the addition of multiple test repetitions or increased load may be necessary to ensure paraspinal and hip extensor fatigue during the Sørensen test. The short rest period that we employed between test repetitions likely resulted in cumulative and progressive muscle fatigue across test repetitions.^39^

We found that the lumbar paraspinals fatigued at a similar rate in the two groups. This supports earlier studies investigating young, or minimally disabled individuals with LBP.^16,18,40^ Compared with older, more disabled individuals, physical deconditioning and disuse atrophy are less likely to have occurred in young adults and in our study total and vigorous physical activity was not significantly different between the individuals with LBP and the back-healthy controls. The shorter Sørensen test duration in the individuals with LBP was also not explained by elevated fear avoidance beliefs. The lack of significant association between fear avoidance and test duration in the present study may be due to the small range of FABQ scores observed in this study. The average score in the physical domain (9.2) was below the threshold for clinically relevant elevated fear avoidance.^41^ In addition, neither pain experienced following the Sørensen test repetitions in either group, nor average pain reported during symptomatic episodes influenced performance on the final repetition of the test. Although post-test pain differed between the groups following the first and second Sørensen repetitions, this group difference was smaller than the minimal detectable difference for visual analogue scales for back pain.^42^

We found that the factors that predicted endurance in the third Sørensen test varied across groups. In back-healthy individuals, lumbar paraspinal fatigability during the third test predicted duration of that final test. This is consistent with previous work in healthy individuals demonstrating a significant linear relationship between the median frequency slope of the lumbar paraspinals and the duration of the Sørensen test.^4,18,43^ However, this relationship between lumbar paraspinal fatigability and test duration was not evident in the young adults with LBP. In the individuals with LBP, the only measure of muscle fatigability that significantly predicted final test duration was the slope of the hamstrings during the first repetition of the test. Similarly, in a sample of individuals with chronic and disabling LBP, Moffroid et al.,^21^ demonstrated a significant correlation between the median frequency slope of the biceps femoris and Sørensen test duration. Our study extends these previous findings to young adults who were minimally disabled and asymptomatic at the time of testing. We speculate that in the individuals with LBP, the fatigability of the hamstrings is more predictive of Sørensen test duration than the fatigability of the lumbar paraspinals because these individuals avoid maximally fatiguing the lumbar paraspinals in order to limit sense of effort in the symptomatic area. Pitcher et al.,^5^ have also reported that fatigability of the biceps femoris during the Sørensen test differed significantly between individuals with and without LBP but that this difference was only evident when additional loading was added to the upper trunk. During sustained sub-maximal contractions such as the Sørensen test, EMG amplitude of muscle activation increases over time as firing frequency and number of recruited motor units increases to maintain force production.^44^ In our participants with a history of LBP, the extent to which they activated the lumbar and thoracic paraspinals was more important than the fatigability of the paraspinal muscle groups in predicting test duration.

Our findings challenge the usual interpretation of the Sørensen test as a measure of lumbar paraspinal fatigability. In comparison with the pain-free group, our participants with a history of LBP used a coordination strategy that was more dependent upon the thoracic and hip extensors. As activation and fatigability of the lumbar paraspinals did not differ between groups, we theorize that in individuals with LBP, greater than normal activation of the lumbar musculature would be needed to compensate for the peripheral changes in lumbar paraspinal structure that are associated with LBP^45^, and that without this compensation endurance during the Sørensen test is impaired. Our study also demonstrates that the difficulty of the Sørensen test must be increased, via the addition of extra trunk loading or test repetitions, to fully identify impairments in young adults with LBP.

We acknowledge some limitations to this study. The PAS is validated against other measures of activity and cardiovascular fitness. However, like all subjective measures it may represent an over-estimation of activity.^46^ In addition, the use of surface electromyography did not enable this study to differentiate between components of the paraspinal muscle group.^16,47^ The amount and distribution of mass contained in the head, arms and trunk vary across individuals. Therefore, the torque that individuals must exert to maintain a horizontal position will be greater in individuals with larger mass in the trunk^23^, which was not quantified or controlled for in this study. However, we did not observe a significant relationship between duration of the third test and BMI.

## Conclusion

In young, active adults, more than one repetition of the Sørensen test is needed to adequately assess paraspinal and hip extensor muscle endurance. Whereas in young adults without symptoms, fatigability of the lumbar paraspinals contributes most to test duration, young adults with LBP, even between symptomatic episodes, use a coordination strategy that involves greater reliance on the thoracic paraspinals and hip extensors.

## Data Availability

All data produced in the present study are available upon reasonable request to the authors.

